# Needle-Plug/Piston-Based Modular Mesoscopic Design Paradigm Coupled with Microfluidic Device for Large-scale Point-of-Care Pooled Testing

**DOI:** 10.1101/2024.06.09.24308631

**Authors:** Baobao Lin, Bao Li, Wu Zeng, Yulan Zhao, Huiping Li, Yin Gu, Peng Liu

**Affiliations:** Department of Biomedical Engineering, School of Medicine, Tsinghua University, Beijing, China; Changping Laboratory, Beijing, P. R. China; State Key Laboratory of Space Medicine, China Astronaut Research and Training Center, Beijing,100094, China

**Keywords:** SARS-CoV-2, Microfluidic, Point-of-care testing, Modular design

## Abstract

Emerging diagnostic scenarios, such as population surveillance by pooled testing and on-site rapid diagnosis, highlight the importance of advanced microfluidic systems for in vitro diagnostics. However, the widespread adoption of microfluidic technology faces challenges due to the lack of standardized design paradigms, posing difficulties in managing macro-micro fluidic interfaces, reagent storage, and complex macrofluidic operations. This paper introduces a novel modular-based mesoscopic design paradigm, featuring a core “needle-plug/piston” structure with versatile variants for complex fluidic operations. These structures can be easily coupled with various microfluidic platforms to achieve truly self-contained microsystems. Incorporated into a “3D extensible” design architecture, the mesoscopic design meets the demands of function integration, macrofluid manipulations, and flexible throughputs for point-of-care nucleic acid testing. Using this approach, we developed an ultra-sensitive nucleic acid detection system with a limit of detection of 10 copies of SARS-CoV-2 per mL. This system efficiently conducts large-scale pooled testing from 50 pharyngeal swabs in a tube with an uncompromised sensitivity, enabling a truly “sample-in-answer-out” microsystem with exceptional performance.

## Introduction

Sudden outbreaks of large-scale human infectious diseases, exemplified by the COVID-19 pandemic^1–4^, present a formidable challenge to public health, social and economic stability, and global security. Early, rapid, and accurate diagnosis of infectious diseases is essential for the effective containment of disease spread and improved treatment outcomes. While conventional laboratory-based qPCR methodology played a vital role in the fight against COVID-19^5–7^, the emerging diagnostic scenarios, such as population surveillance by pooled testing^8,9^, on-site rapid diagnosis^10,11^, and at-home self-testing^12,13^, obviously cannot be fulfilled by the qPCR alone. For example, in pooled testing, the dilution-induced false negative is inevitable when using conventional qPCR. Therefore, a more sensitive system is highly demanded for mixing more samples in a single test without sacrificing the sensitivity and the specificity. Both the on-site and the at-home analyses require instruments that have the true “sample-in-answer-out” capability, which contradicts the laboratory-based qPCR method. Microfluidics has the potential to revolutionize disease diagnosis, offering low-cost, portable, and automated solutions for pathogen detection, especially in the setting of point-of-care testing (POCT)^14–17^. Nowadays various microfluidic platforms, such as centrifugal discs^18,19^, electrowetting-on-dielectric (EWOD)^20,21^, digital droplets^22,23^, etc., have already been developed to enable complex biochemical analyses in a micro-nano scale, including the COVID-19 diagnosis. However, the absolute dominance of the conventional qPCR highlights the necessity of the further improvement of microfluidic systems, especially for these emerging demands.

Almost all the advantages provided by microfluidic systems stem from one major characteristic: miniaturization. The reduction of the processing and reaction volumes can remarkably enhance the analytical efficiency. Moreover, the miniaturization makes it feasible to integrate the entire analytical process on a single device, leading to automation and portability. Unfortunately, when facing the actual needs of *in vitro* diagnostics and many other biochemical analyses, miniaturization may also bring troubles to microfluidics. Clinical samples often come as diverse formats in a broad range of volumes up to several milliliters. Due to the low abundance of detection targets, the processing of large-volume samples is deemed inevitable. In addition, although the reagent consumption in a microfluidic system can be as little as nanoliters, the long-term storage and the accurate, bubble-free loading of the reagents into the microdevice requires much more amount, even beyond the microliter scale.

The volume gap between the macrofluids of samples and reagents and the microstructures poses a significant challenge to the development of fully integrated microdevices for IVD, which may have to sacrifice certain performances. For example, many microfluidic systems require additional manual operations for pre-processing samples and loading reagents prior to the on-chip analysis^24,25^. Alternatively, some microfluidic IVD products were coupled with external liquid handling robots for loading samples and reagents, resulting in bulky footprints of the instruments and the openness to the ambient^26^. Aluminum pouches were also employed to store reagents on-chip^27^. However, it is difficult to assemble these separated pouches in a microdevice and the release of the reagents has a high rate of failure. Overall, the development of standardized, systematic structures that can be seamlessly integrated with various microdevices for handling macroscopic solutions is an urgent task to improve the performance of microfluidic systems.

In the current study, we addressed the critical need to bridge the gap between the macroscopic fluids and microscale structures by developing a novel modular-based mesoscopic design paradigm for integration with microfluidic devices. Inspired by the classical syringe structure, we designed a core structure of the “needle-plug/piston” and a series of variants, which can be sequentially combined to form more complex unit operations for storing, mixing, and releasing solutions in the μL-mL scale. These structures can be easily coupled with various microfluidic platforms to achieve truly self-contained microsystems. We further incorporated this mesoscopic design paradigm into a “3D extensible” architecture^28,29^ proposed previously by our group to fulfill the needs of function integrations, macrofluid manipulations, and flexible throughputs for point-of-care nucleic acid testing. Based on this design method, we then developed an ultra-sensitive nucleic acid detection system with a detection limit of 10 copies of SARS-CoV-2 per mL, which is capable of performing a large-scale pooled testing of 50 pharyngeal swabs in a tube. Such a mesoscopic design paradigm will enable the truly “sample-in-answer-out” microsystem with exceptional performances.

## Results

### The core element of the mesoscopic design paradigm

The core element of the mesoscopic design paradigm is the “needle-plug/piston” structure, consisting of a hollow needle stuck and glued in the reservoir of the microfluidic channel on a planar microdevice as a chip connector and a hollow barrel sealed with a rubber plug from the bottom and a movable rubber piston on the top as a container. A well fixture is usually attached to the upper surface of the planar microdevice for positioning the container in place (Fig. 1a). In the initial position, the needle is half-inserted into the rubber plug of the container, so that the container is fixed in the well fixture while the solution can still be sealed. The container can also be easily loaded into the fixture prior to use if the solution must be refrigerated separately. A steel plunger driven by a stepper motor is employed to actuate this “needle-plug/piston” structure. The container is first pushed downwards and the needle is penetrated through the plug to form a classical syringe configuration for dispensing the solution into the microdevice. Once the solution is fully dispensed, the needle is further inserted into the rubber piston to close the connector completely.

**Fig. 1.**
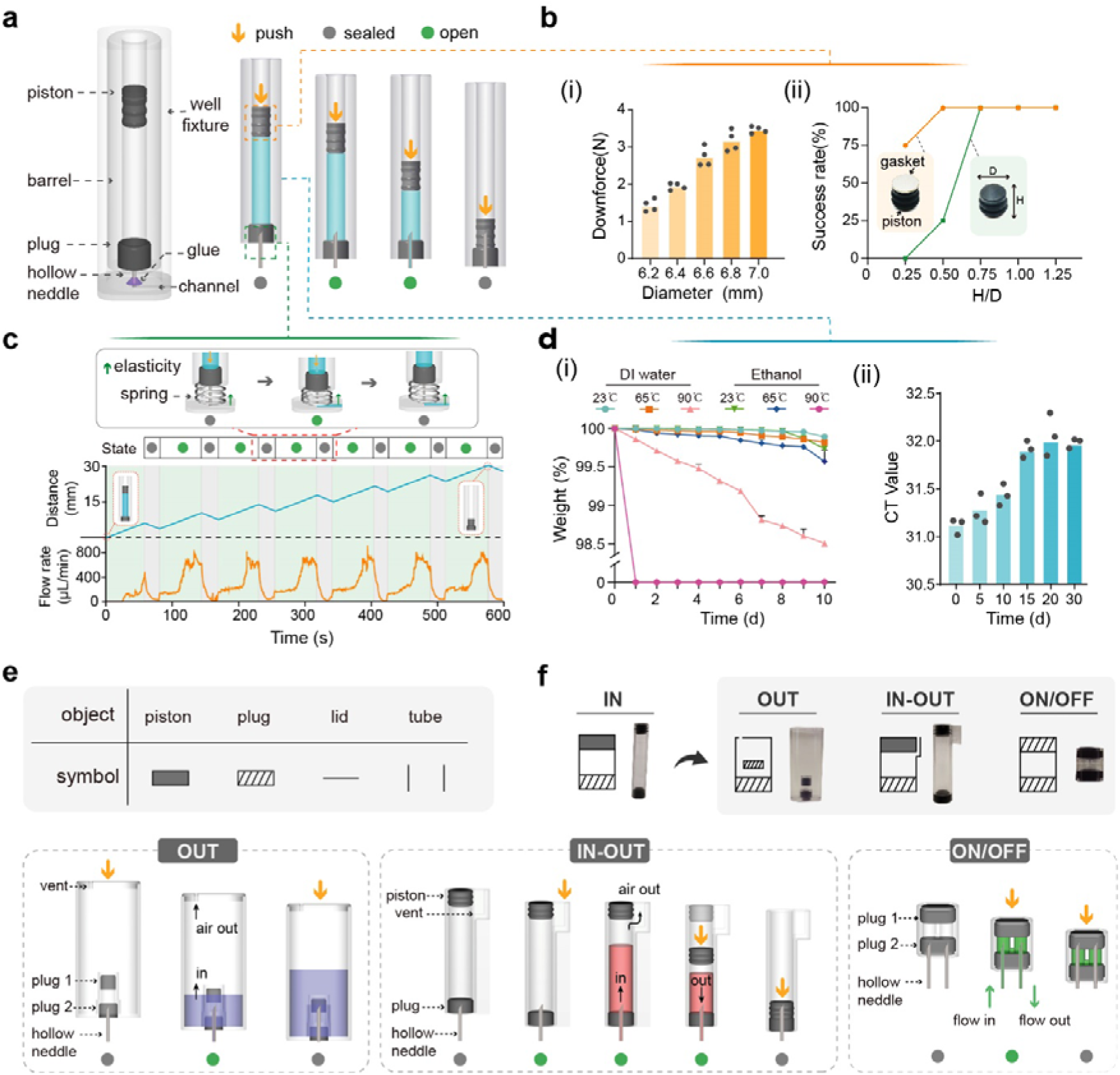
The mesoscopic design paradigm. **a,** Structure and operation of the core “needle-plug/piston” element. The core element undergoes a sequence of sealed-open-sealed states as the piston descends, maintaining an exclusive connection with microfluidic channels during fluid release. **b,** Optimization of piston diameter and H/D (heigh/diameter) ratio. (i) Downforce remains below 4 N with various piston diameter in the same barrel. (ii) The effect of the piston H/D ratio and the use of a gasket on the success rate during the piston actuation process. **c,** Coordinated injection cycles with a spring. Coordinating the core component with a spring enables repeated on-off injection cycles, ensuring consistent fluid release volume and flow rate without leakage. **d,** Container tightness and storage time influence. (i) Sealing tests of containers filled with deionized water and ethanol. Error bars represent mean ± s.d. (n = 3). (ii) Influence of storage time on the biological activity of reaction reagents in containers. PCR premix was stored in containers for 30 days. Every 5 days, the PCR mix was tested for amplification. **e,** Symbolic representation of the elemental objects. **f,** Structural and operational principles of core element variants. OUT element features a double-plug structure, often functioning as a waste container. IN-OUT element includes a shoulder at the top and a vent hole, enabling both the introduction and withdrawal of reagents. ON/OFF element is a valve with two needles and plugs, opening on the first press-down and closing on the second.

We comprehensively characterized this basic structure from the following aspects: first, the accuracy of solution dispensing can be adjusted by tuning the diameter of the barrel and the step of the stepper motor. Considering the fabrication of the barrel, the alignment of the plunger to the piston, and potential tilting issues during the fixing needle process (Extended Data Fig. 1), we estimated the minimum diameter of the barrel is about 3 mm, which results in a minimum dispensing volume of 0.7 μL. Second, we measured the force required for the piston actuation and found the maximum force is about 4 N. In addition, by adding a steel gasket on the top of the piston and increasing the depth-to-width ratio of the piston, the stability of the piston movement can be significantly improved (Fig. 1b). Third, the extensive testing of various thicknesses and hardness of the rubber plug as well as the diameters and the bevel angles of the hollow needle demonstrated that the maximum force required for penetrating through the plug is less than 7N (Extended Data Fig. 2). When only a part of the solution is injected, a spring can be installed in the bottom of the container. The retraction of the plunger results in the pushback of the container and the seal-off of the needle with the rubber plug. This on-off cycle can be repeated without any leakage of the solution (Fig. 1c and Extended Data Fig. 3).

The design of the container is like a serum vial, the bottom of which is just replaced with a rubber piston. As a result, the air-free filling of the containers can be adapted to the mature vial filling lines. The long-term storage of solutions was tested with deionized water and ethanol (Fig. 1d (i)) in the containers injection-molded with polypropylene (PP). After stored at room temperature and 65°C for 10 days, the losses of the reagents were both less than 0.4%. At a high temperature of 90 °C, ethanol slowly dissolved PP, resulting in leakage, while the loss of DI water is still under 1.5%. Furthermore, we found a PCR mix stored in the container for 30 days can still provide a similar bioactivity to the original (ΔCt < 1) (Fig. 1d (ii)). In addition, the container can be produced with glass for the storage of corrosive solutions. Overall, this “needle-plug/piston” structure provides an ideal means for operating and storing liquids.

### Various basic elements for basic fluidic operations

The core element described above represents one of the basic fluidic operations, i.e., loading a reagent into the microdevice, which is defined as IN. Based on this core structure, we developed a set of basic elements for various basic operations, including OUT, IN-OUT, and ON/OFF, by changing the configuration of the needles, plugs, and pistons in the container. For the ease of drawing a schematic, all these elements are assigned with symbols designed with a wireframe system (Fig. 1e, f). The OUT element means driving a solution out of the microdevice and its function is achieved by adding a double-plug structure on the bottom of the container. In addition, the piston of the container is replaced with a vent hole, which can be covered with a piece of hydrophobic film to prevent the leakage of the solution and aerosols. The first pushdown of the container lets the needle penetrate through the first plug. At this position, the solution in the microchannel can be driven into the container. After that, the container is pushed down again to let the needle be sealed with the second plug. The OUT element usually functions as a waste container in the microdevice. The IN-OUT element has a shoulder on the top of the container. The plunger first pushes on the shoulder to let the entire container move downwards, so that the needle penetrates through the plug. After that, the solution in the microchannel can be driven into the container, and the air in the container is vented through a vent hole. After that, the vent can be closed by pushing down the piston. If the piston is further pushed down, the solution can be loaded into the microchannel again. The ON/OFF element has two needles and two plugs without the barrel to only function as a valve. The first press-down of the structure by the plunger can open the valve and the second closes it.

### Modular-based mesoscopic design paradigm for fluid operations

For the ease of the development of microdevices with this mesoscopic design paradigm, we developed a set of validated unit operations of macrofluids which can be combined and thereby realize any application-specific assays on the platform. These unit operations are realized by sequentially linking the above-mentioned basic elements via the microchannels in the microdevice. First, multiple stock solutions are often sequentially loaded into a microchip for downstream reactions and analyses. A sequential injector was designed to accomplish this operation by linearly linking multiple IN elements as illustrated in Fig. 2a (i) and demonstrated in Supplementary Video 1. Due to the seal of each container by either the plug or the piston, the solutions can be sequentially loaded into the channel by simply pushing down the containers one by one without the worry of unintentionally mixing solutions in the containers. Second, the liquid distribution is another common operation (Fig. 2a (ii) and Supplementary Video 2). By linking an IN element with multiple OUT elements, we can distribute the solution in the IN container to the following several containers. Third, for the transportation of liquids, fluid valving is often needed to control the flow path. As shown in Fig. 2a (iii) and demonstrated in Supplementary Video 3, the fluidic valving can be easily achieved by adding ON/OFF elements between the IN and OUTs. Fourth, we can accomplish the mixing of diverse reagents by combining multiple INs and IN-OUT, as illustrated in Fig. 2a (iv) and demonstrated in Supplementary Video 4. The mixing of two reagents within the IN-OUT is achieved through the uniform dispersion of bubbles, subsequently leading to further reactions in subsequent steps. These verified unit operations form a component library, which can used for integration with various microfluidic platforms.

**Fig. 2.**
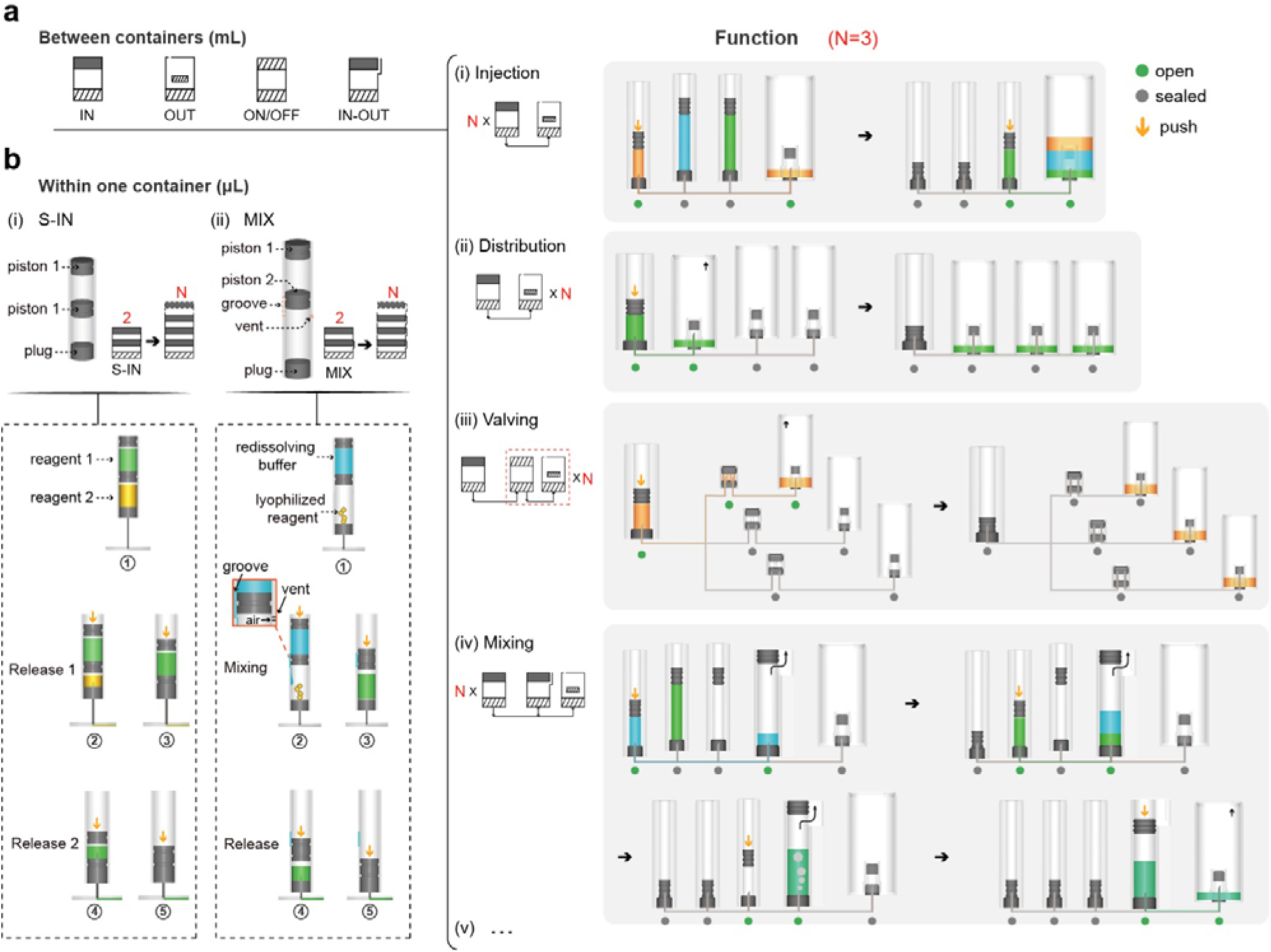
Modular-based mesoscopic design paradigm for fluid operations. **a,** Versatile element combinations for macro-scale liquid manipulations (mL) among containers: (i) Injection: linking multiple IN elements with an OUT element; (ii) Distribution: linking an IN element with multiple OUT elements; (iii) Valving: adding ON/OFF elements between the IN and OUTs; (iv) Mixing: combining multiple INs and IN-OUT. **b,** Fluid manipulations (μL) within one container. (i) Structure and operational principles of the multi-release element (S-IN). The S-IN features a hollow barrel with multiple pistons isolating various reagents. Applying downward pressure to the top piston sequentially connects the hollow needle at the bottom with each reagent, facilitating their release. (ii) Structure and operational principles of the multi-mix element (MIX). The MIX consists of lyophilized reagents in the lower layer and redissolving buffer in the upper layer. Applying downward pressure to the top piston allows the redissolving buffer to enter the lower layer through grooves on the surface of the barrel. Air from the lower layer is expelled through the vent, facilitating effective mixing. The mixed reagent is then released for subsequent reactions.

To achieve a compact design without using too many containers, more complex fluid manipulation can be achieved within a single container by incorporating multiple pistons and by carving grooves and holes on the surface of the barrel. Here we demonstrated two liquid handling components, which we defined as advanced elements. First, the multi-release element consists of a hollow barrel and multiple pistons (Fig. 2b (i) and Supplementary Video 5). These pistons separate the container into multiple segments, which can be used to store different reagents (S-IN). When the top piston is pushed down by the plunger, the hollow needle on the bottom punctures through each piston, leading to the release of the reagents into the device sequentially. This advanced element is suitable for storing several reagents in a single container to reduce the footprint of the structures. Second, the multi-mix element is also composed of multiple pistons to separate a container into several segments (Fig. 2b (ii) and Supplementary Video 6). By milling grooves on the outer surface of the barrel and drilling holes, the segments in the container are connected. Unlike the multi-release element, the redissolving buffer in the top segment of the element is first pushed down to mix with the lyophilized reagents in the second segment. Eventually, all the reagents are mixed together and then driven into the microfluidic channel.

### Integration with various microfluidic platforms

Our mesoscopic design paradigm provides a flexible and standardized means of macrofluidic operations for various microfluidic systems. We streamlined the design process into the following three steps (Fig. 3a): 1) to identify the interface and functionalities required to connect the microfluidic platform with macroscopic reagents; 2) to glue hollow needles at the designated interface, serving as a connection point for the mesoscopic components; 3) to attach a well fixture to the upper surface of the microfluidic platform and to insert the corresponding components.

**Fig. 3.**
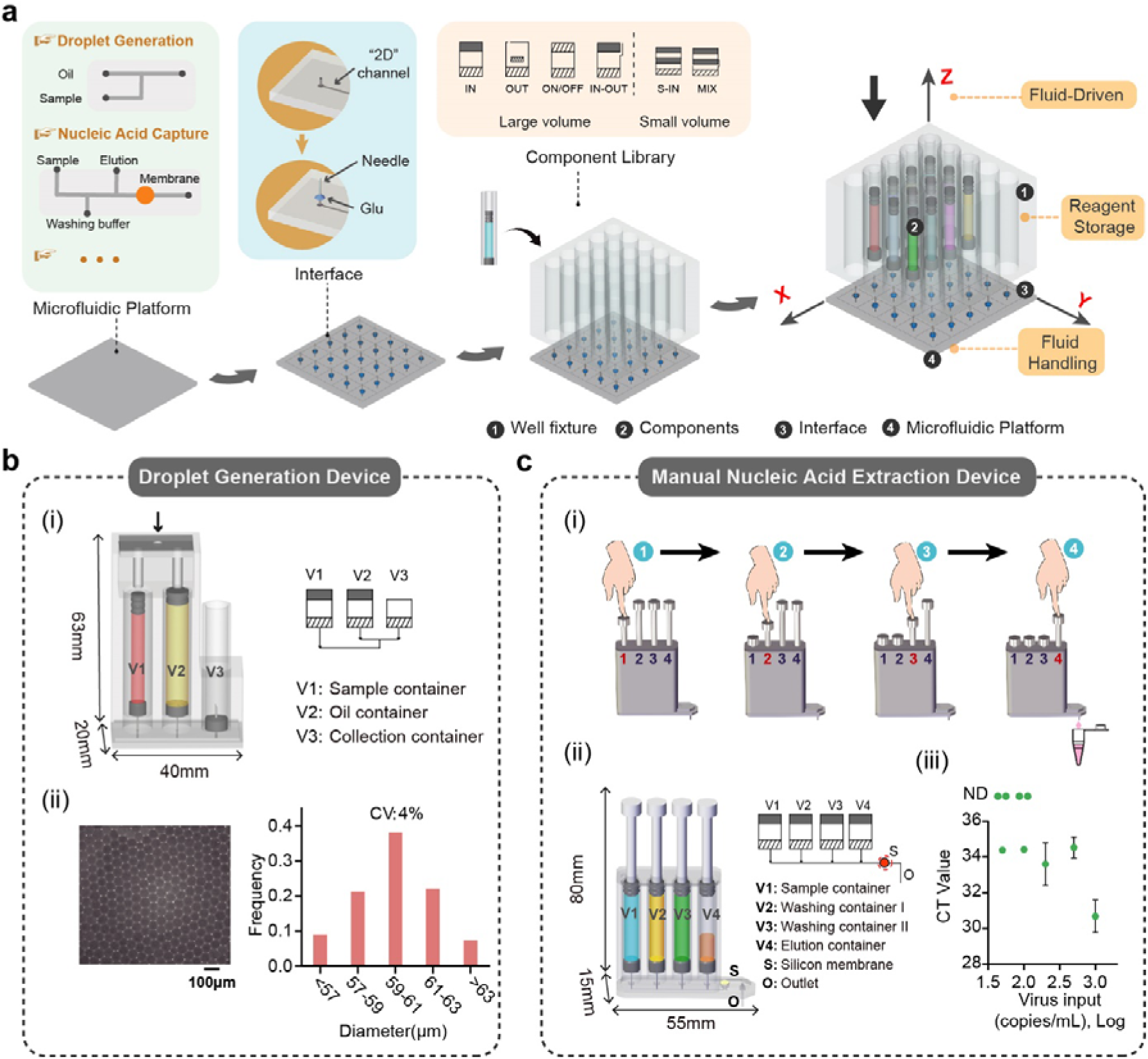
Design guidelines for seamless integration with diverse microfluidic platforms. **a,** Three-step integration of mesoscopic layer structures with microfluidic platforms: 1) To identify the interface and the functionalities for connecting macroscopic reagents; 2) To glue hollow needles for macroscopic components; 3) To attach a well fixture and insert corresponding containers. In the integrated system, fluid-driven power is provided from the top through a plunger. Components within the system are designed for reagent storage and macroscale manipulations, and the lower microfluidic platform optimizes the connection of different components, facilitating fluidic handling and reactions. **b,** Droplet generation device. (i) Structure and operational principles. V1 contains the aqueous phase and V2 contains the oil phase, both of which are connected to the ends of a T-shaped channel. The droplet generation process utilizes a diameter ratio of D1:D2=1:3 between V1 and V2. Simultaneously pressing down the pistons results in the oil-phase flow at 450 microliters/hour and the water flow at a speed of 150 microliters/hour. (ii) Visualization and particle size distribution of the generated droplets. **c,** Manual nucleic acid extraction device. (i) Procedure for operating the manual nucleic acid extraction device. (ii) Structure of the device. V1 to V4 are IN elements for sequentially injecting the sample, the washing buffer I, the washing buffer II, and the elution buffer through a silicone membrane. (iii) Sensitivity test of SARS-CoV-2 virus extractions using the manual nucleic acid extraction device. Error bars represent mean ± s.d. (n = 3).

As an example, we showcase the integration of our mesoscopic structures with a typical microfluidic device for droplet generation, which is a key step for many biochemical assays, such as digital PCR^30,31^, single-cell analysis^32,33^, etc. Although the generated droplet in the microchannels is in the nanoliter-volume scale, the total reagent consumption can be up to milliliters. In addition, the long-term storage of oil also poses a significant challenge to the system design. As a result, many digital PCR systems still require the manual reagent loading process prior to the on-chip operations. Here we demonstrated that a T-shaped droplet generation chip can be easily transformed into a stand-alone device using our mesoscopic design paradigm (Fig. 3b). Two IN containers for the sample and the oil phase, respectively, and one collection container for collecting droplets can be attached to the planer microchip, on which needles are installed as the connectors. Simultaneously pressing down both the pistons of the containers using the plunger allows precise flow rate control and the collection of the generated droplets in the collection container. We conducted tests on the size and distribution of the generated droplets. Despite a coefficient of variation (CV) of 4%, which falls short of the ideal, further optimization is achievable by fine-tuning the channel size and speed. Notably, with this additional mesoscopic interface layer, the classical droplet generator becomes a truly enclosed, self-contained system.

Another advantage of our modular-based mesoscopic design paradigm lies in the actuation mode, i.e. all types of containers can be actuated by a simple downward press, which is carried out using either an automated stepper motor or just a finger. This manual operation feature is extremely important to the emerging scenario of at-home self-testing, which is more sensitive to the initial instrument cost. Here we designed a manual nucleic acid extraction device, in which a piece of silica gel membrane is embedded for DNA/RNA capture (Fig. 3c). By sequentially pressing the four buttons of the device, the sample, washing solution I, washing solution II, and elution are sequentially driven through the silica gel membrane and the nucleic acid extraction from the sample is achieved. We tested that the extracted nucleic acid solution from the manual device provides a sensitivity of 200 copies/mL of the SARS-CoV-2 RNA, with an optimal elution volume of 8 microliters (Extended Data Fig. 4).

### “3D extensible” architecture for the development of an ultra-sensitive microdevice for COVID-19

In the quest to develop fully integrated microdevices for point-of-care testing, we are facing several critical challenges, including the increasing complexity of assays, the volume gap between the macroscale samples and the microstructures, and the diverse throughput demands in various clinical settings. Previously, we proposed a “3D extensible” design architecture^28,29^, in which the function integration, the “world-to-chip” interface, and the adjustable throughput were fulfilled in the X, Y, and Z directions, respectively. Here we integrate the modular-based mesoscopic design structures with the “3D extensible” architecture. As illustrated in Fig. 4a, in the slim cassette-shaped device, the flow direction in the device is defined as the X axle (the “function” direction), along which the elements presented above are linked via the channels in the bottom chip to perform a complete bioassay. In the vertical Y axle (the “interface” direction), all the elements are actuated by the top plunger and the heights of the containers can be adjusted according to the sample and reagent volumes. The cassette can be arrayed along the Z direction (the “throughput” direction) to achieve a flexible throughput. The number of cassettes is adjustable to meet the throughput need of each run.

**Fig. 4.**
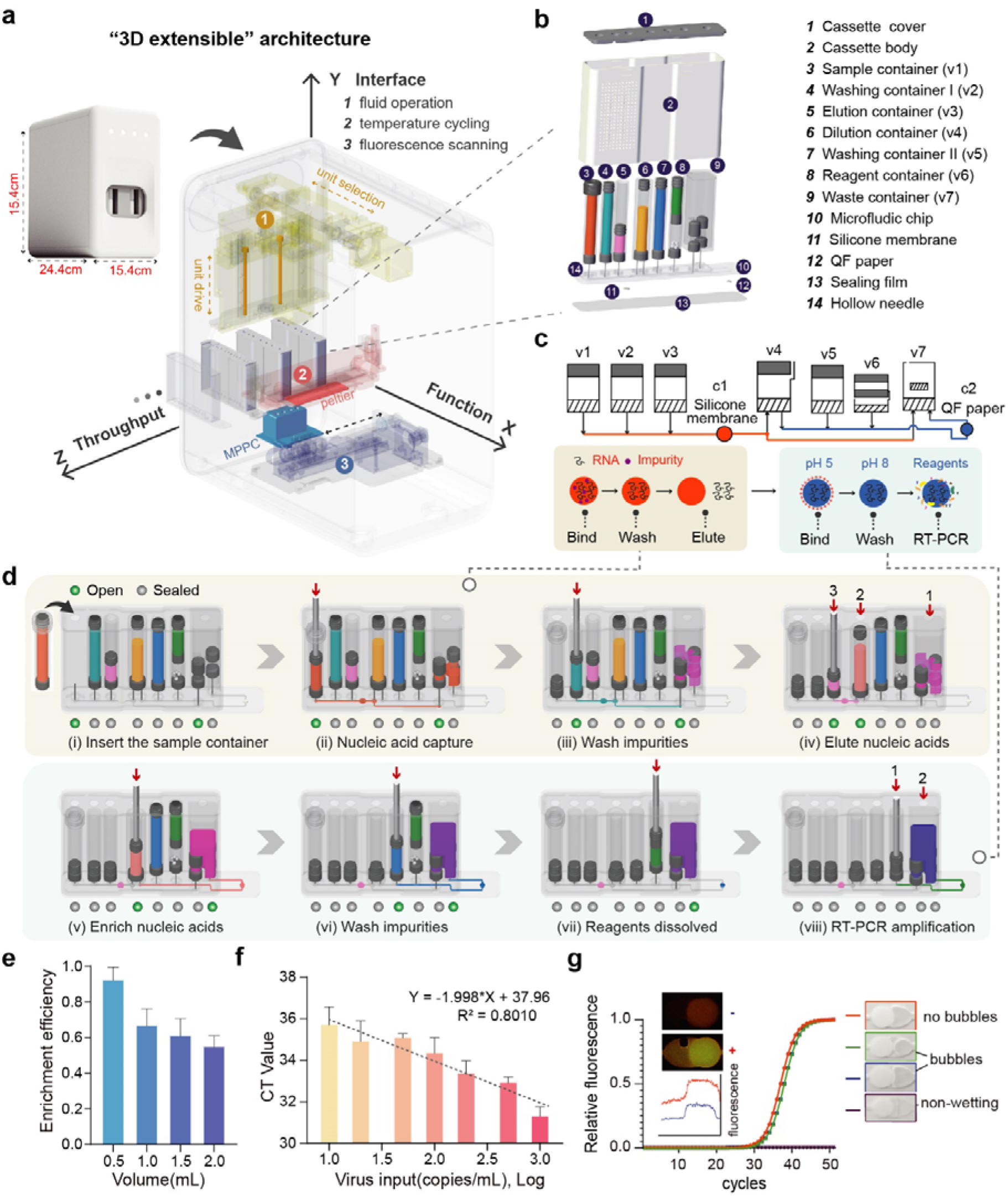
Fully integrated double-extraction-qPCR microdevice (iDEP) in a “3D extensible” architecture. **a,** Schematic of the “3D extensible” architecture design in a cassette-shaped device. In a cassette-shaped device, flow along the X-axis (“function” direction) connects elements through channels, enabling the function integration. The Y-axis (“interface” direction) regulates element actuation with adjustable container heights based on volumes. Cassettes can be arranged along the Z-axis (“throughput” direction) for flexible throughput. **b,** Schematic representation of the cassette structure. All reagents are pre-stored in the cassette, allowing for room-temperature storage. **c,** Principle of the double-extraction method. Nucleic acids are initially extracted from lysed samples using a silica membrane. The eluted nucleic acid solution from the silica membrane undergoes secondary enrichment using the QF paper. **d,** Fluid schematic of the cassette. (i)-(iv) illustrates the nucleic acid extraction process, facilitating the impurity removal from the sample using a silica membrane. (v)-(viii) demonstrates the secondary enrichment of nucleic acids, followed by in situ amplification. **e,** RNA capture efficiency of QF paper. The QF paper is fixed within the cassette chamber, and the sample is laterally passed through it. Various quantities of SARS-CoV-2 RNA templates were introduced to the QF paper. Following a washing step, the QF paper was taken out for RT-PCR amplification in PCR tubes. Error bars represent mean ± s.d. (n = 3). **f,** Evaluation of SARS-CoV-2 RNA detection sensitivity using the double-extraction method in the cassette. Error bars represent mean ± s.d. (n = 3). **g,** In situ nucleic acid amplification using QF paper. The red and blue curves depict the fluorescence intensity of positive and negative amplification results, respectively. To evaluate the impact of bubbles in the reaction, PCR were performed in the chamber under the no-bubble, the small-bubble, the large-bubble, and the non-wetting of the paper conditions.

Pooled testing has been proven to be a cost-effective way to quickly identify infected individuals and prevent them from spreading the disease during the COVID-19 pandemic. In pooled testing, due to the inevitable target dilution induced by the pooling, at most 5-10 samples are grouped and tested together using the conventional qPCR method. A more sensitive methodology is highly demanded for mixing more samples in a single test without sacrificing the sensitivity. Previously, we have developed an integrated microfluidic cartridge that utilizes a chitosan-modified quartz filter (QF) paper^34^ and “*in situ*” tetra-primer recombinase polymerase amplification (tpRPA)^35^ for detecting SARS-CoV-2 in aerosols. Due to the elimination of the elution step, the sensitivity of this integrated microdevice can be improved to 20 copies/mL SARS-CoV-2. However, we found this sample enrichment method has a limited capability of dealing with samples containing many impurities, such as pooled samples. Therefore, we envision that we can first employ a conventional silica membrane for extracting nucleic acids from raw samples. Subsequently, the nucleic acids eluted from the silica membrane undergo a secondary enrichment using the above-mentioned QF paper (Fig. 4c). This double-extraction method not only efficiently removes any impurities, but also ensures the near 100% use of captured nucleic acids.

After the determination of the entire analytical process, we developed a fully integrated double-extraction-qPCR microdevice (iDEP) (Fig. 4b) for ultra-sensitive pooled testing using the “needle-plug/piston” mesoscopic design paradigm under the “3D extensible” architecture. In this design, the first-step capture chamber (c1) containing a piece of silica membrane and the second *in situ* capture and amplification chamber (c2) containing a piece of QF paper are first designed on the planar microdevice. Three containers (v1-v3) functioning as the sample loading, the wash, and the elution steps are connected to the c1 chamber, while another three containers (v4-v6) used as the sample conditioning, the wash, and the reagent loading steps are linked to the c2. Both the chambers share the same waste container (v7), which can be switched to open to c1 and c2 sequentially.

The operation of this device is achieved by constructing a modular-based control and detection instrument, which contains a 2-dimensional stepper motor module for chip actuation, a Peltier-based thermal cycling module for RT-PCR, and a fluorescence linear scanning module for detection (Extended Data Fig. 5-9). The operation of the iDEP device comprises eight steps (Fig. 4d and Supplementary Video 7). Firstly, the sample loading can be achieved by simply inserting the container into the device without the direct handling of the sample (i). The next three steps are the typical capture-wash-elute procedure for the first-round nucleic acid extraction (ii-iv). After that, the secondary enrichment of the eluted nucleic acids followed by the solubilization and the loading of RT-PCR reagents for in situ amplification is realized in the subsequent three steps (v-vii). Finally, the RT-PCR is carried out in the chamber (viii).

### Performance characterization of the iDEP device

We first evaluated the extraction efficiencies of the silica membrane and the capture efficiencies of QF paper using the standard SARS-CoV-2 RNA (Fig. 4e and Extended Data Fig. 10). The nucleic acid extraction efficiency of the silica membrane decreases with an increase in nucleic acid concentration. At a concentration of 500 copies/mL, it can achieve a capture efficiency of approximately 85%, maintaining around 70% efficiency at higher nucleic acid concentrations. In the context of QF paper, the capture efficiency exhibited a gradual decline from nearly 100% to 50% with an increase of the sample volume from 0.5 to 2 mL. In the iDEP device, the nucleic acid extracted through the silica membrane undergoes the pH adjustment with a volume of less than 200 μL before passing through the QF paper. As a result, nearly all the nucleic acids extracted by the silica membrane can be effectively captured by the QF paper. Moreover, both the silica membrane and the QF paper demonstrate high efficiencies at lower virus RNA concentrations, thereby enabling the ultra-high sensitivity. Subsequent tests revealed that the iDEP can provide a limit of detection (LOD) of 10 copies of SARS-CoV-2 RNA per mL, significantly surpassing most of detection methods (Fig. 4f). Other than the high sensitivity, another advantage provided by the *in situ* amplification is the tolerance for bubbles during the PCR in the reaction chamber. We introduced various degrees of bubbles into the amplification chamber during the reagent loading on purpose. We found that the amplification primarily occurs on the QF paper. As long as the filter paper is soaked in the PCR mixture, the amplification remains completely unaffected by the bubbles in the chamber, making the PCR more robust (Fig. 4g).

The operation of this iDEP system is as simple as the following: insert the sample container into the cassette, load the cassette into the instrument, and press the start button. The entire testing process takes only 45 minutes, and the reagents are pre-stored in the cassette, enabling room-temperature storage. This versatility makes it suitable for various application scenarios (Fig. 5a and Extended Data Fig. 11). We collected pharyngeal swabs from 10 individuals, introducing pseudo-viruses at different concentrations to simulate various virus levels. The results show consistent detection for different individuals at 20-50 copies/mL (Fig. 5b), an order of magnitude higher than the standard method (400-500 copies/mL). Moreover, we conducted an assessment experiment of our fully integrated nucleic acid detection system with pharyngeal swab samples collected from 76 individuals. Both the cassette and the standard magnetic bead-based extraction method were employed for nucleic acid detection (Fig. 5c). The results demonstrate that the cassette effectively detected both N and ORF1ab genes of SARS-CoV-2 in 38 positive samples, surpassing the limitations of the traditional magnetic bead-based RT-qPCR method in certain cases. These findings emphasize the superior detection sensitivity of our system in clinical applications. Both methods yielded negative results for 38 negative samples, confirming the excellent detection specificity of our system. A comparative analysis with qPCR instrument Ct values revealed a robust linear correlation (Extended Data Fig. 12), suggesting that our system achieves relative quantification of viral nucleic acid, with the Ct value reflecting virus concentration. These results not only validate that our system meets clinical testing requirements but also highlight its practical value in application.

**Fig. 5.**
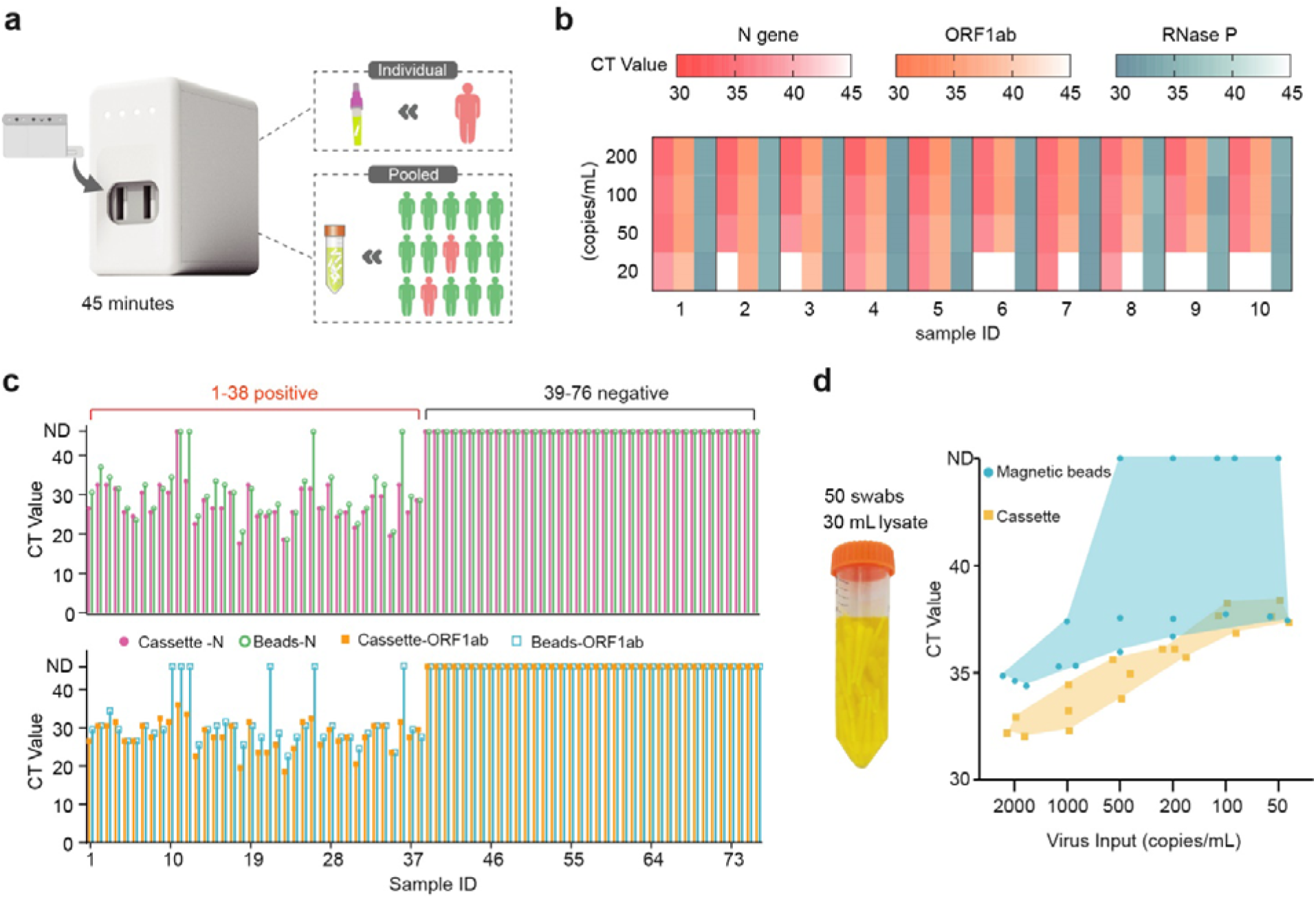
Performance characterization of the iDEP device. **a,** Fully integrated nucleic acid analysis system (iDEP). The iDEP system facilitates the testing of individual and pooled samples within 45 minutes in a sample-in-answer-out manner. **b,** Evaluation of the detection sensitivity for single pharyngeal swab samples. Ten swabs from healthy individuals were tested by adding various pseudo-viruses to assess the system’s sensitivity. **c,** Detection results of 76 clinical samples. These samples were parallelly tested using the iDEP system and the conventional RT-qPCR method. **d,** Evaluation of the detection sensitivity in large-scale pooled testing. A total of 50 pharyngeal swabs from 50 healthy individuals are collected in in a tube with 30 mL of lysis buffer. Different concentrations of pseudo-viruses were added in the lysis buffer and tested using both the iDEP system and the conventional magnetic bead-based nucleic acid extraction with RT-qPCR.

### Large-scale pooled testing performed by the iDEP device

The ultra-high sensitivity of our iDEP system should be beneficial for large-scale pooled testing. In the testing of a single-person sample, a pharyngeal or nasopharyngeal swab is typically inserted into a 3-mL collection solution. By contrast, in a typical pooled testing, 10 swabs are collected in a 10-mL collection solution. As a result, to maintain the same sensitivity and specificity, the pooled testing should be at least 3.3 times more sensitive and 3 times more capable of removing impurities than those of the single-sample testing. As we demonstrated above, our iDEP system can provide a limit of detection of 20-50 copies/mL, which is about 10 times more sensitive than the conventional method. Therefore, according to the above calculation, we reason that a large-scale pooled testing with around 50 swabs could be tested using the iDEP. To test this hypothesis, we first found at least 30 mL of solution is required to soak 50 pharyngeal swabs (Fig. 5d). Next, we introduced pseudo-virus templates at various concentrations into the 30-mL lysis buffer with 50 swabs and tested using our iDEP system and the conventional qPCR with magnetic bead-based nucleic acid extraction. The results show that the iDEP maintains a stable detection limit at 50 copies/mL (equivalent to 500 copies/mL in a single-person sample). In contrast, traditional nucleic acid testing is limited to 500 copies/mL (equivalent to 5000 copies/mL in a single-person sample), leading to potential false negative results. This underscores the significant advantages of our iDEP in the large-scale pooled testing, ensuring effective detection and substantial savings in reagent costs and time.

## Discussion

While the microfluidic technology takes advantage of miniaturization to realize the integration, automation, and high efficiency of bioassays, the volume gap between the macroscale solutions and the microscale structures often results in the sacrifice of certain performance of the integrated microsystems, such as lengthy manual sample preparation, bulky instrument, low sensitivity, etc. To remedy this gap using a standardized and versatile solution, we developed a modular-based mesoscopic design paradigm to function as additional layers attached to any microfluidic systems for dealing with large-volume-scale samples and reagents. There are several critical advantages provided by this design platform. First, the core “needle-plug/piston” element together with the variant structures enables the reliable storing and actuating of samples and reagents in the volume range from several microliters to milliliters. In this structure, the plug/needle interface can provide a secure, reversible, and convenient connection between the mesoscopic layer and the microchip without any solution leakage. The container can be separately manufactured and stored, providing flexibility in mass production and use. Moreover, a pre-validated library of unit operations can be constructed with these basic containers to provide a standardized solution for common macrofluidic manipulations, expediting the development of new microsystems.

Second, the fluid actuation of one-way pressing simplifies the instrument and even requires no instrument at all. Although the upward movement of the piston in a container can be realized to achieve more complex liquid manipulation, we only employed the downward pressing of the pistons on purpose. The one-way actuation can make the structures of the external instrument as simple as possible and the loading of the device into the instrument can be easily conducted without the requirement of the precise alignment. Moreover, like the manual nucleic acid extractor demonstrated above, even no instrument can be realized with our design platform for the application, where the initial instrument cost is a big concern, such as at-home self-testing.

Third, the integration of the modular-based mesoscopic paradigm with the “3D extensible” architecture provides a reliable platform for the development of point-of-care molecular diagnostic microsystems. Based on this architecture, a slim cassette-like microdevice can be developed to address the need for complex function integration, microfluidic manipulation, and flexible throughput. This unified structure not only expedites the development process but also lowers the costs of the devices by using many common parts.

As a demonstration, we have designed a fully integrated, ultra-sensitive microsystem for detecting the SARS-CoV-2 virus. By implementing a dual nucleic acid extraction method in our cassette, we achieved a remarkable detection sensitivity as low as 10 copies/mL. The system successfully tested 76 clinical pharyngeal swab samples, producing results consistent with the standard qPCR method and underscoring its clinical significance. Furthermore, this system can be employed for large-scale pooled testing, ensuring the same sensitivity (500 copies/mL) as that of non-pooled testing while also reducing testing costs and time. For example, envision airport customs where testing all individuals on an airplane could be achieved with just a few samples.

In summary, our mesoscopic design paradigm addresses the challenges of reagent storage, sample pretreatment, and complicated large-volume solution manipulations that are difficult to realize merely using microfluidic platforms. By integrating with other microfluidic technologies, our platform enables the development of a more complex and fully integrated system without sacrificing performance in a short development process at a lower expense.

## Methods

### Microfluidic elements and cassettes fabrication

All microfluidic elements and cassettes were designed using AutoCAD 2019 and SolidWorks 2018 software for 2D and 3D drawings. All the container bodies of the microfluidic elements are injection molded with polypropylene (PP), and the plugs and the pistons are injection molded with butyl rubber.

The droplet generation device and the manual nucleic acid extraction device are precision-manufactured using polymethyl methacrylate (PMMA). The iDEP device for SARS-CoV-2 detection is injection molded with polycarbonates (PC), and the main body and upper cover of the cassette are injection molded with acrylonitrile butadiene styrene (ABS). The pressure-sensitive adhesive tape is cut into shape using a cutting die. The assembly of the cassette consists of four main steps: i) installing the hollow needles; ii) fixing the internal cassette body; iii) assembling the microfluidic components; and iv) installing the cassette shell.

### Storage validation of reagents

Sealing tests were conducted to assess the storage conditions of reagents in the container. First, 700 μL of deionized water and 700 μL of ethanol were loaded in the containers, which were then sealed with a plug at the bottom and a piston at the top. The containers were stored at 23LJ, 65LJ, and 90LJ, and their weights were recorded every day. For the biological activity test of the reagent storage, a PCR mixture containing RT-PCR mix, primers, and probes for the N gene of SARS-CoV-2 was stored in containers at −20LJ. Every 5 days, the reagents in 3 containers were tested for RT-qPCR.

### Design and control of the iDEP analyzer

The home-built compact iDEP analyzer contains three modules: a fluid driving module, a thermal cycling module, and a fluorescence scanning module (Extended Data Fig. 5-9). Each module is designed with AutoCAD 2019 and SolidWorks 2018 software for 2D and 3D drawings and manufactured with machining. All the modules are synchronously controlled via Modbus by a laptop installed with a home-made LabVIEW software (Extended Data Fig. 11).

The analyzer is set into operation by initiating the fluid driving module. The sample in the sample container (v1) is first driven at a speed of 800 μL/min through a silicone membrane, followed by the washing buffer in the washing container I (v2) at a speed of 500 μL/min. Subsequently, the nucleic acid extraction pathway of waste container (v7) is closed, and the dilution container (v4) is opened. Then, the elution buffer in the elution container (v3) is driven at a speed of 100 μL/min to facilitate the mixing with the pH adjustment buffer in the dilution container (v4). After the first extraction process, the acidic elute in the dilution container (v4) is driven at a speed of 100 μL/min through the QF paper, followed by the loading of DI water in the washing container II (v5) at a speed of 500 μL/min. After the loading of the reagent in the reagent container (v5) at a speed of 50 μL/min, the waste container (v7) is completely closed and the thermal cycling begins. The thermal cycle consists of a reverse transcription step at 52LJ for 300 seconds, followed by activation at 95LJ for 15 seconds. The subsequent 45 cycles include denaturation at 95LJ for 10 seconds and annealing/extension at 60LJ for 35 seconds. The optical detection is performed during the annealing/extension step by the fluorescence scanning module.

### Mechanical testing of needle penetration through plug

All the hollow needles are manufactured by Shenzhen Zhuding Metal Products Co., Ltd (China). All the rubber plugs used for the initial testing are made by 3D printing. The mechanical testing equipment is purchased from Bengbu Dayang Sensing System Engineering Co., Ltd (China). The needle is fixed on the mechanical sensor by a needle holder, and the force sensor records the force changes as the needle is slowly pressed into the rubber plug (Extended Data Fig. 13).

### Fluid testing platform

The structure of the fluid testing platform is shown in Extended Data Fig. 14, which includes a Z-axis motor and an X-axis motor. The Z-axis motor drives the push plunger to move up and down to drive microfluidic components, while the X-axis motor drives the push plunger to move left and right to select different microfluidic components. These two motors are controlled by a controller, which can configure the distance and speed of the operation and can also run automatically according to the set program. The microfluidic cassette is placed on a platform and fixed in its initial position by a fixture in the left-right and front-back directions. During the fluid driving process, a camera is used to capture real-time images at the bottom of the platform, which is displayed on a laptop connected to the platform. All microfluidic components, component combinations, and fluid tests of the microfluidic cassette are conducted on this platform.

### Droplet generation device

Reagents were loaded into the droplet generation device as follows: the sample container contained 150 μL of aqueous phase reagent, the oil container contained 500 μL of oil (purchased from TargetingOne Co., Ltd.), and the collection container was empty. The plunger simultaneously presses down the sample container and the oil container, generating droplets. Both the sample and oil containers are IN components. Once the droplets are generated, they are examined under a microscope, and the resulting images are processed using ImageJ to obtain statistical data on the particle size distribution.

### Manual nucleic acid extraction device

Reagents loaded into the manual nucleic acid extraction device are as follows: 700 μL of lysis buffer in the sample container, 700 μL of washing buffer I in the washing container I, 700 μL of washing buffer II in the washing container II, and 50 μL elution buffer in the elution container, all of which were purchased from Nanjing Vazyme Biotech Co., Ltd. (RC313-01). Silicon membrane was purchased from ABigen Co., Ltd. (109265A011). In the experiment for volume optimization of the eluted template solution in the amplification reaction, 200 copies of the SARS-CoV-2 template were added to the sample container. A 50-microliter eluate was obtained using the manual nucleic acid extraction kit, and different volumes of the eluent containing the template were subsequently added to 25 microliters. RT-qPCR detection was performed in the amplification system to determine the optimal eluted template solution volume.

### Integrated double-extraction-qPCR microdevice (iDEP)

All the containers used in the device are first cleaned with pure water, followed by sterilization and mold removal treatment. Reagents loaded into the microfluidic cassette for SARS-CoV-2 detection are as follows: 1-1.2 mL of lysis buffer (purchased from Nanjing Vazyme Biotech Co., Ltd.)in the sample container, 700-800 μL of alcohol-containing washing buffer (purchased from Nanjing Vazyme Biotech Co., Ltd.) in the wash container I, 100 μL of RNase-free ddH2O in the elution container, 600 μL of 10 mM MES (2-Morpholinoethanesulphonic acid) buffer at pH=5.0 in the dilution container, 700-800 μL of deionized (DI) water in the wash container II, and 100 μL of DI water and lyophilized PCR mix pellets (purchased from Zhuhai Bao Rui Biotechnology Co., Ltd.) in the reagent container.

### Modification of quartz filter paper with chitosan

Reagents for the filter paper modification includes low-molecular-weight chitosan, MES (2-(N-morpholino) ethanesulfonic acid), SDS (sodium dodecyl sulfonate), and GPTMS ((3-glycidyloxypropyl) trimethoxysilane), all of which were purchased from Sigma-Aldrich (St. Louis, MO). The quartz filter paper was from Whatman (QHA, GE Healthcare, Pittsburgh, PA). All solutions were prepared in water purified to 18.2 MΩ cm by Milli-Q Advantage A10 (Millipore, Massachusetts, MA). The chitosan modification process involved treating a 47 mm diameter piece of quartz filter with GPTMS (2.5% in methanol) for 1 hour to add epoxy groups on the surfaces of filter fibers. Subsequently, the treated filter was immersed in a 20 mL MES solution containing 10 μL of 1% chitosan (prepared with 0.1% acetic acid, pH 6.0) and incubated overnight on a shaker. After modification, the filter paper was washed with acetic acid (0.1%, pH 6.0) three times and dried completely at 50 °C in a vacuum drying oven. 2 mm diameter discs of the quartz filter paper were then punched out and stored in a sealed Petri dish at room temperature until use.

### Performance verification of the QF paper

To assess the RNA capture efficiency of the QF paper, various volumes of lysis solution containing pseudovirus (5000 copies/ml) were introduced into sample containers. These sample containers were then inserted into a microfluidic cassette and placed on the fluid testing platform for driving. Subsequently, the microfluidic cassette was disassembled and the paper was subjected to RT-qPCR amplification. The One-step PrimeScript RT-PCR (TaKaRa) kit was used for nucleic acid amplification. The composition and temperature cycling of the RT-PCR mix used in this study, unless otherwise specified, were as follows: Each reaction contained 25 μL of RT-PCR mixed solution, 0.5 μM of forward primer, 0.5 μM of reverse primer, and 0.3 μM of probe. The thermal cycling protocol involved a reverse transcription step at 52 °C for 6 min, followed by an initial activation at 95 °C for 10 s, and then 45 cycles of denaturation at 95 °C for 5 s and annealing/extension at 60 °C for 30 s.

For in-situ amplification experiments using QF paper, the amplification results of the positive and negative groups were captured as fluorescent images using a dual LED blue/white light transilluminator (Solarbio). The obtained images were processed using ImageJ software to obtain the distribution data of their fluorescent intensities. To investigate the effect of bubbles on amplification, we used 1 ml of the viral template with a concentration of 1000 copies/mL of lysate. The sample was passed through filter paper at a flow rate of 800 μL/min, followed by passing through 700 μL of washing solution at a flow rate of 500 μL/min. When adding the RT-PCR mix, we controlled the volume of the liquid injected into the reaction chamber to regulate the formation of bubbles in the chamber.

### Standard RT–PCR test for pharyngeal swab samples

The nucleic acid extraction in the standard RT-PCR detection process was conducted using the GeneRotex fully automated nucleic acid extraction instrument (Xi’an Tianlong Technology Co., Ltd.), while the RT-PCR was performed on the Applied Biosystems 7500 Real-Time PCR Instrument. For pharyngeal swab sample detection, the collected samples were partitioned into two groups and tested using the standard RT-PCR detection process and the developed fully integrated detection system. The Ct values for the N gene and the ORF1ab gene were recorded to enable the comparison of the two methods. The primer sequences used in this study are listed in Extended Data Table 1.

### Detection of SARS-CoV-2 pharyngeal swab samples

76 samples collected in the experiment were from symptomatic patients or individuals in close contact with patients. All clinical pharyngeal swab samples were obtained with approval from the Tsinghua University

Institutional Review Board (THU01-20230093).

## Author contributions

B. Lin and B. Li designed and performed experiments, analyzed the data and prepared the manuscript. P.L. conceived the project, designed experiments, provided overall supervision for this work and wrote the manuscript. Y.G. provided the QF paper for in situ amplification and co-supervised this work. W.Z. helped design the microfluidic cassettes and diagnostic instrument. Y.Z. and H.L. performed diagnostic RT-qPCR testing of the respiratory swab samples.

## Data Availability

All data produced in the present study are available upon reasonable request to the authors

## Acknowledgements

This work was financially supported by the National Key Research and Development Program of China (Grant No. 2022YFC2704902) and the National Natural Science Foundation of China (Grant No. 32001021).

