## supplemental file for "Needle-Plug/Piston-Based Modular Mesoscopic Design Paradigm Coupled with Microfluidic Device for Large-scale Point-of-Care Pooled Testing"

**This supplementary file includes the following information:**


- Table S1 The sequence of primers used in this study.
- Figure. S1 Key factors affecting barrel diameter.
- Figure. S2 Effect of rubber plug and hollow needle parameters on penetration force.
- Figure. S3 Experimental details of the on-off cycle for reagent injection.
- Figure. S4 Optimization of eluted solution volume.
- Figure. S5 The modular design of the nucleic acid analyzer.
- Figure. S6 Schematic and performance parameters of the fluid driving module.
- Figure. S7 Schematic and performance parameters of the temperature cycling module.
- Figure. S8 The performance validation of the temperature cycling module.
- Figure. S9 Structure and performance of the fluorescence scanning module.
- Figure. S10 The RNA capture efficiency of the silica membrane.
- Figure. S11 The iDEP system.
- Figure. S12 Comparison of CT values obtained through iDEP and conventional qPCR method.
- Figure. S13 Force testing apparatus for puncturing rubber with a needle.
- Figure. S14 Photograph of the fluid testing platform.

**Table S1 The sequence of primers used in this study.**

| Primers | Sequence (5'-3') |
|---|---|
| N-F | GCTTCTGACACAACTGTGTTCAC |
| N-R | CGGCAGACTTCTCCACAGGAGT |
| N-P | 5'-Cy5-ACCTCAAACAGACACCATGG -BHQ1-3' |
| ORF1ab-F | ACGGGTTTGCGGTGTAAGTGCAG |
| ORF1ab-R | AGATGTCAAAAGCCCTGTATACG |
| ORF1ab-P | 5'-FAM-ACACCGTGCGGCACAGGCACTAG-BHQ1-3' |
| RNase P-F | AGATTTGGACCTGCGAGCG |
| RNase P-R | GAGCGGCTGTCTCCACAAGT |
| RNase P-P | 5'-VIC-TTCTGACCTGAAGGCTCTGCGCG-BHQ1-3' |

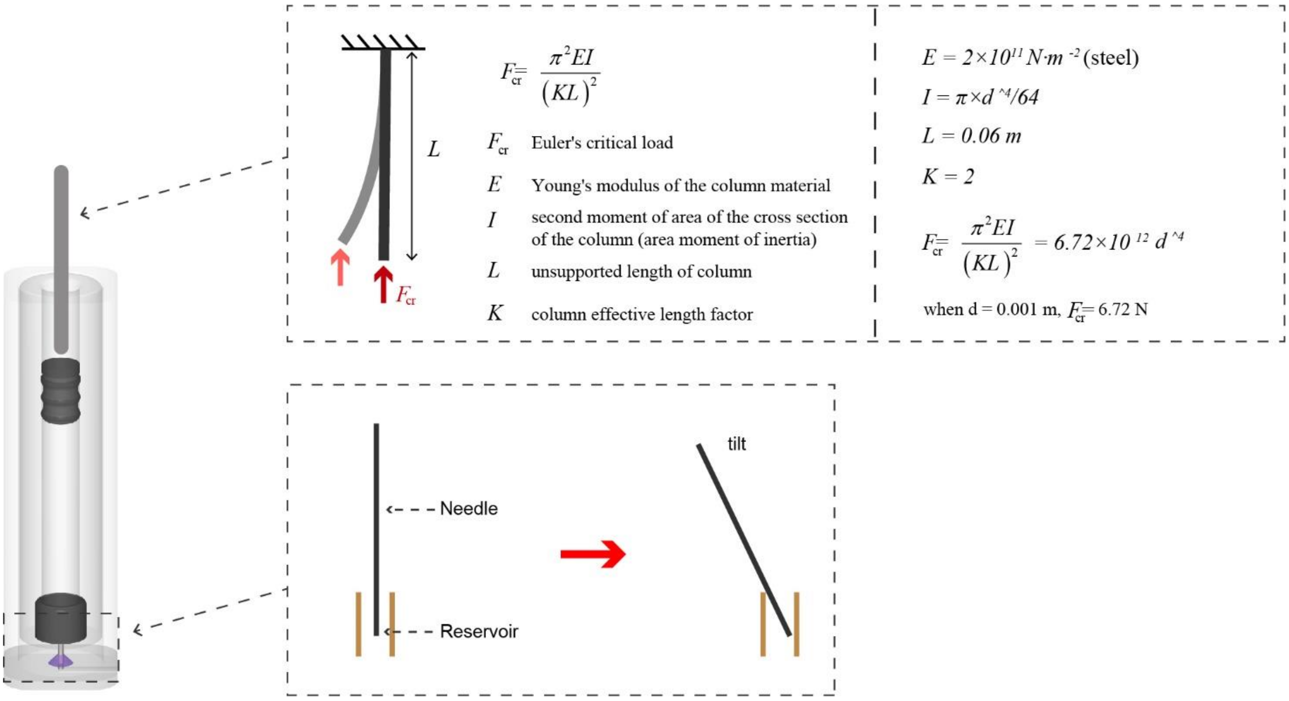


**Figure. S1 Key factors affecting barrel diameter.**

The factors affecting the barrel diameter can be broadly categorized into two types. Firstly, machining-induced errors play a crucial role. To ensure the sealing integrity of the components, the inner diameter of the barrel must not exhibit significant variations. Theoretical calculations indicate that when the plunger is made of steel and has a diameter of 1mm, it can provide a force of 6.72N without deformation during the downward movement, exceeding the force required in our tests when the barrel diameter is 7mm. Theoretically, the minimum barrel diameter can be below 1mm. However, practical challenges arise during production processes, and maintaining consistent barrel diameters, especially in smaller sizes, is difficult, whether through machining or injection molding (due to the larger aspect ratio).

The second type of influencing factor is errors introduced during assembly and operation. In securing the hollow needle in the reservoir, a certain gap is intentionally left between the reservoir and the hollow needle for ease of assembly. This gap may result in needle tilting, necessitating a proportionate increase in the barrel diameter to accommodate these errors. Additionally, errors are inevitable during the alignment of the plunger with the piston, making it challenging to ensure 100% concentricity. Therefore, a reasonable increase in the barrel diameter is necessary. Taking all these factors into consideration, we have opted for a barrel with an inner diameter of 3mm.

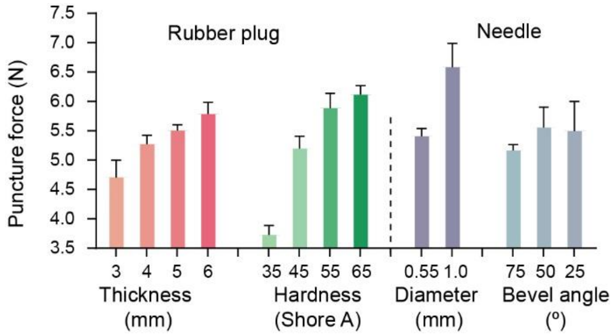


**Figure. S2 Effect of rubber plug and hollow needle parameters on penetration force.** In the rubber plug thickness test, a consistent hardness of 45° was maintained, whereas in the hardness test, a thickness of 3mm was held constant. The diameters test involved keeping a needle bevel angle of 45° constant, while a diameter of 0.55mm was maintained in the bevel angle test. Error bars represent mean ± s.d. (n = 3).

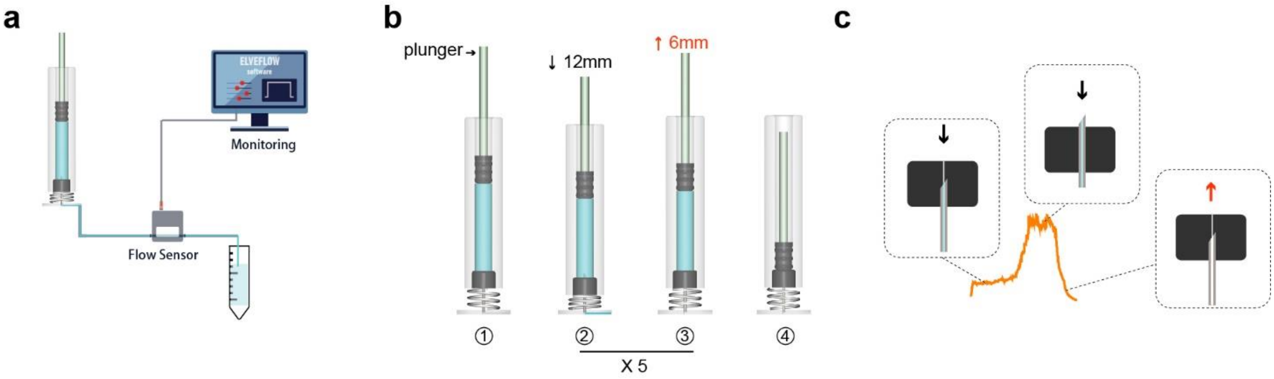


**Figure. S3 Experimental details of the on-off cycle for reagent injection. a,** Connection of the hollow needle's base to the flow sensor (procured from *Elveflow*) enables the measurement of the flow rate for each release of reagent. **b,** Details of the plunger movement throughout the process involve a downward motion of 12mm, followed by an upward movement of 6mm. This cycle repeats five times until all reagents are completely dispensed. **c,** In each cycle, the position relationship between the hollow needle and the rubber plug corresponds to fluid flow rates. As the plunger drives the piston downward, subtle cracks in the plug, resulting from the previous penetration by the hollow needle, allow fluid to exit the container at a relatively low flow rate due to internal pressure. Once the hollow needle fully pierces the plug, the flow rate reaches its maximum. As the plunger moves upward, relieving pressure, the container reseals.

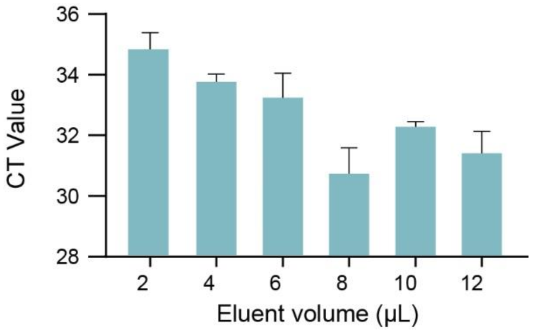


**Figure. S4 Optimization of eluted solution volume.** The experimental reaction volume is 25 μL, with the addition of 1 μL of elution fluid in the reaction system. Error bars represent mean ± s.d. (n = 3).

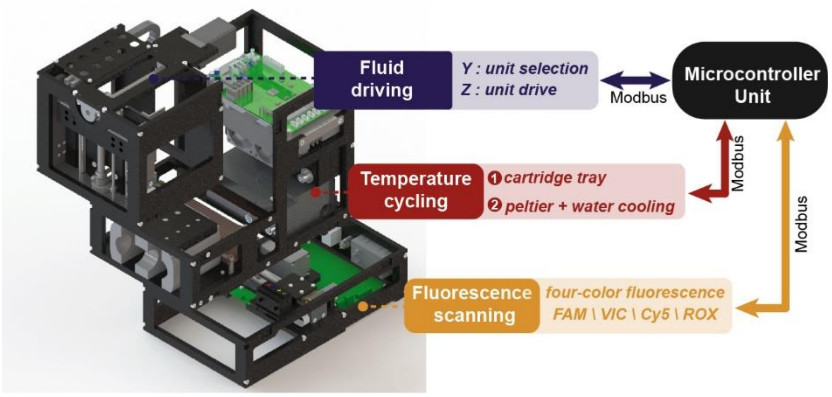


**Figure. S5 The modular design of the nucleic acid analyzer.** The analyzer is divided into three modules: Fluid Driving, Temperature Cycling, and Fluorescence Scanning. These modules are interconnected to the microcontroller unit through the Modbus protocol.

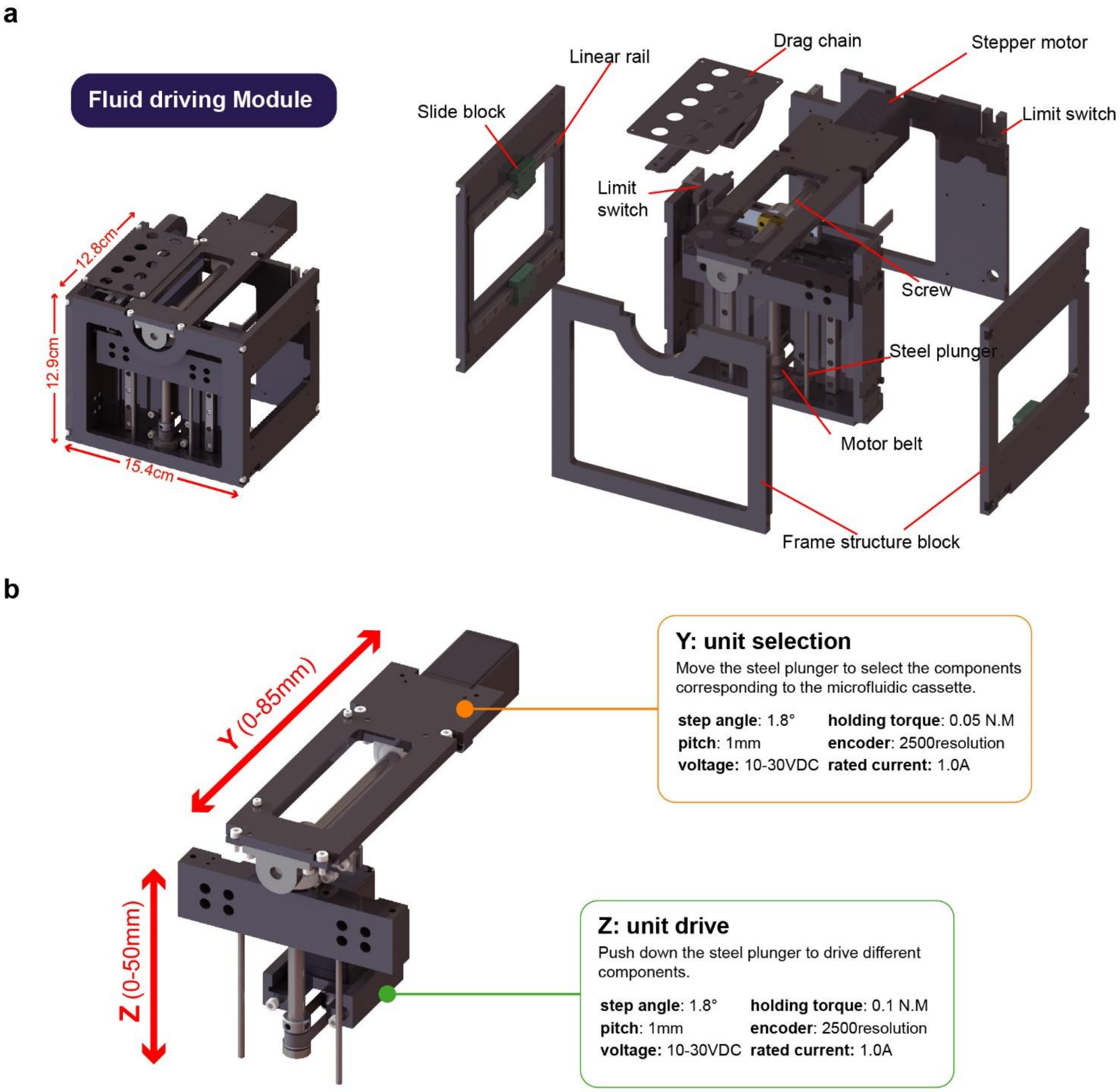


**Figure. S6 Schematic and performance parameters of the fluid driving module. a,** Exploded view of the fluid driving module. **b,** Functional and performance parameters of the Y and Z axes of the fluid driving module.

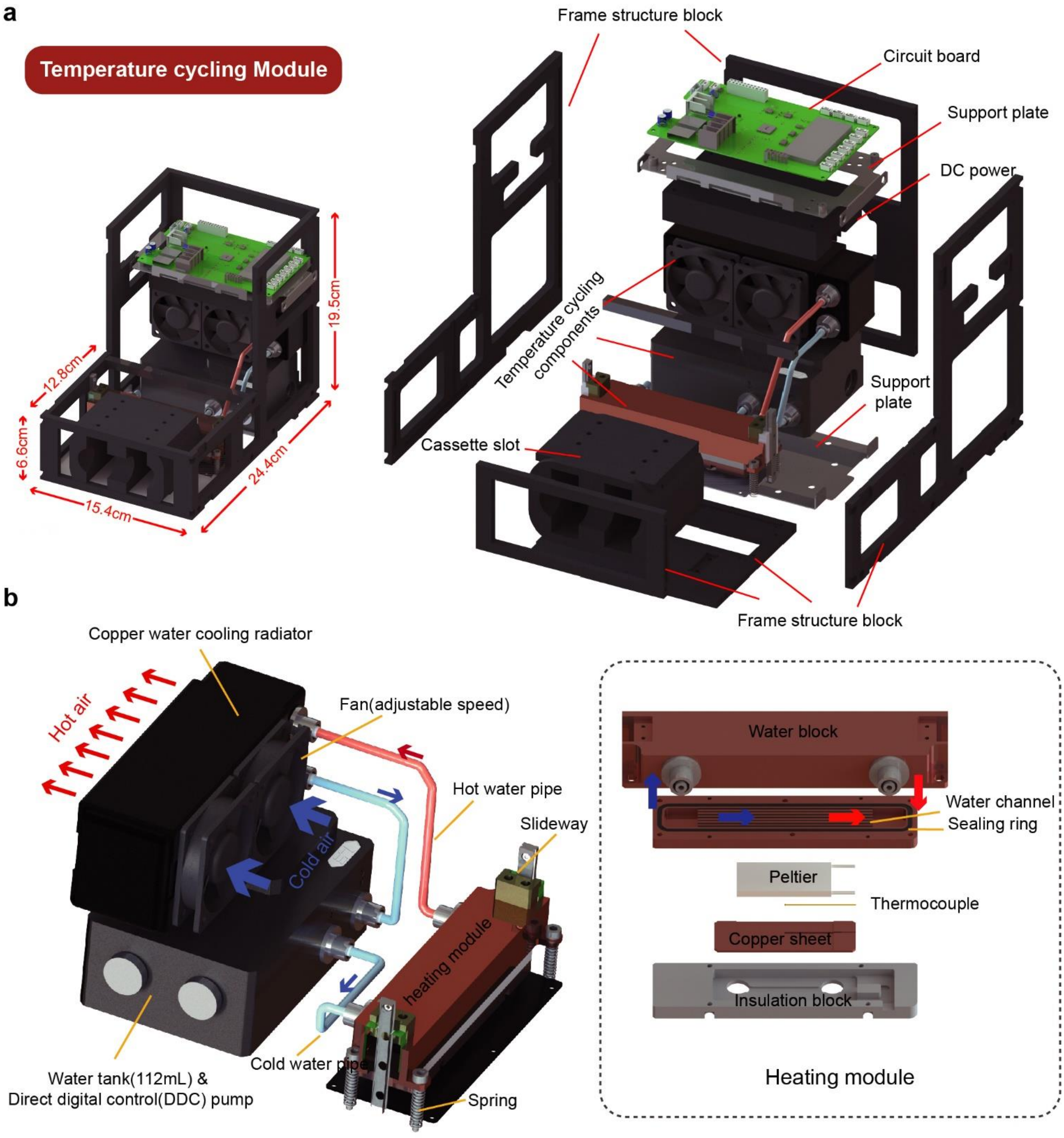


**Figure. S7 Schematic and performance parameters of the temperature cycling module. a,** Exploded view of the temperature cycling module. **b,** Working principle of the temperature cycling module. The module uses Peltier elements for heating and cooling, and a water-cooling system for heat dissipation.

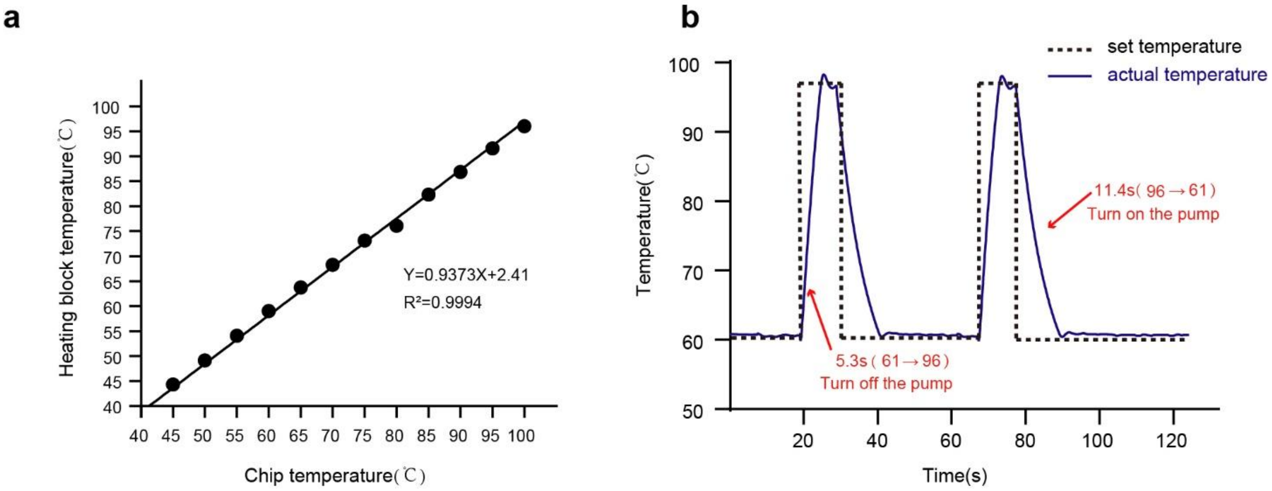


**Figure. S8 The performance validation of the temperature cycling module. a,** Relationship between the temperature inside the chip and the temperature of the heating block. **b,** Temperature curve inside the chip during the PCR temperature cycling process.

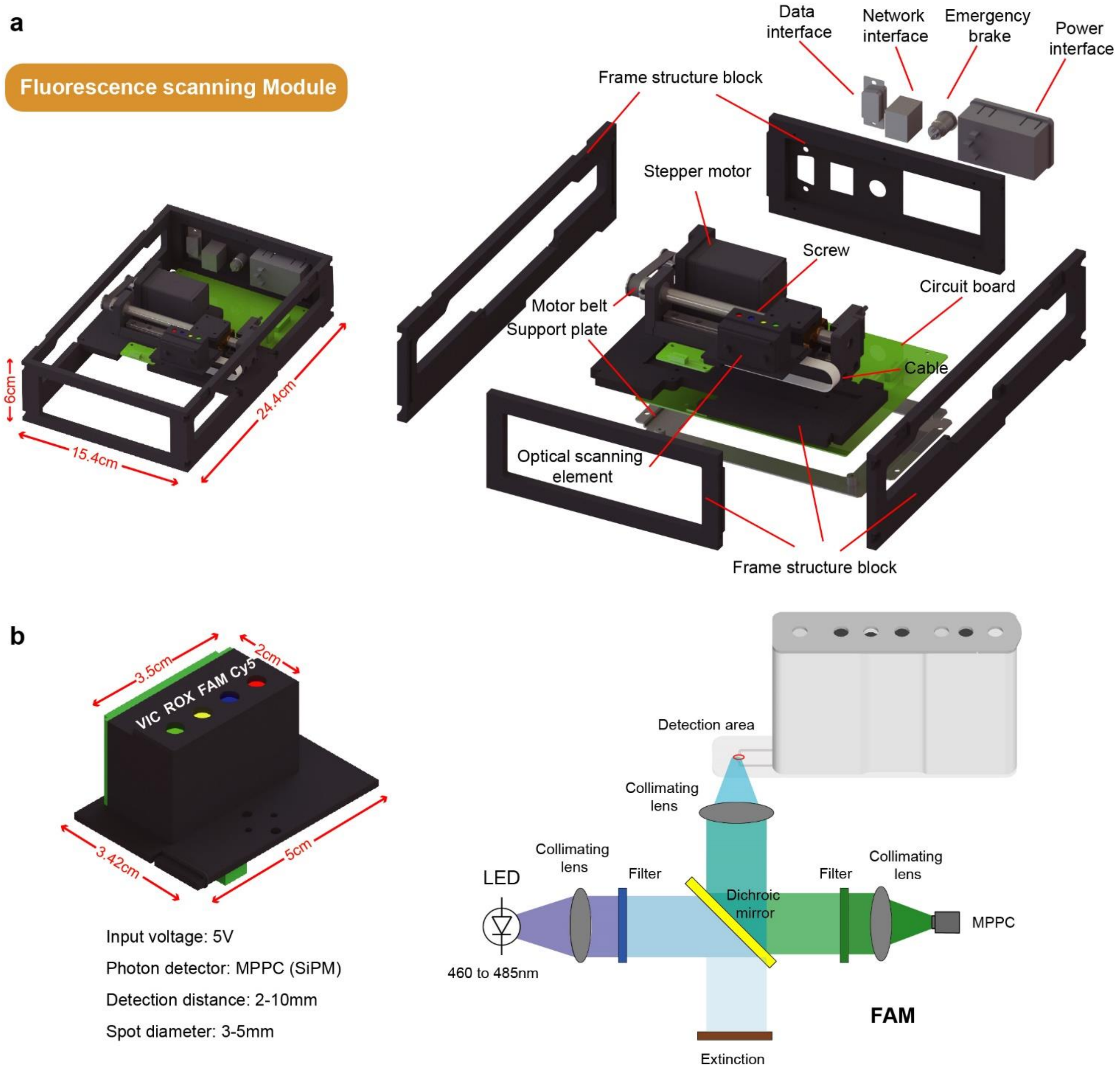


**Figure. S9 Structure and performance of the fluorescence scanning module. a,** Exploded view of the fluorescence scanning module. **b,** Working principle and detection performance of the fluorescence scanning module.

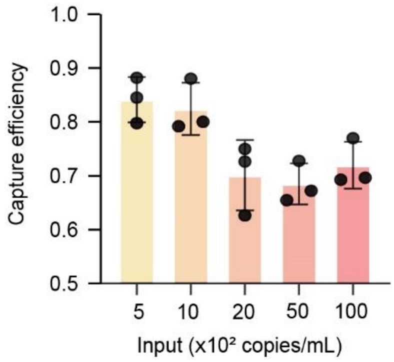


**Figure. S10 The RNA capture efficiency of the silica membrane.** The membrane is fixed within the cassette chamber, and the sample is laterally passed through it. Various quantities of SARS-CoV-2 RNA templates were introduced to the membrane. The elution solution is used for RT-PCR amplification. Error bars represent mean ± s.d. (n = 3).

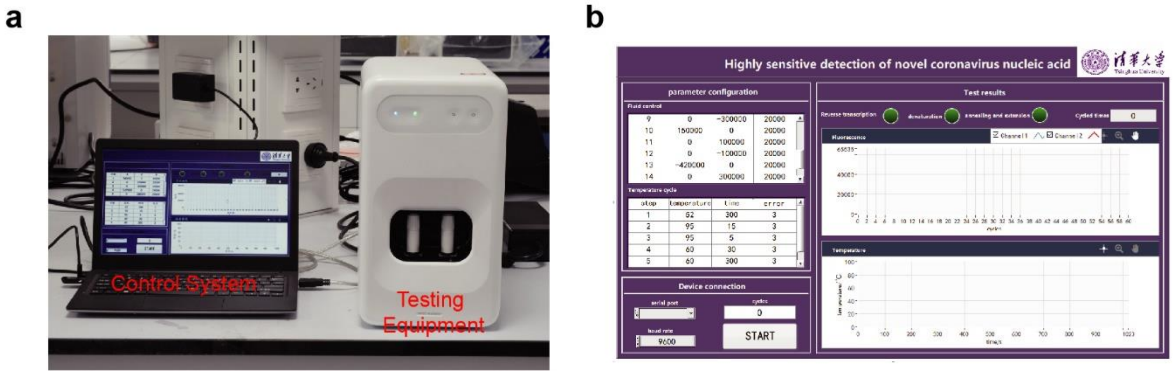


**Figure. S11 The iDEP system. a,** Composition of the fully integrated analyzer. **b,** LabVIEW control interface. The LabVIEW software interface is designed with parameter configuration, equipment connection, and result display sections.

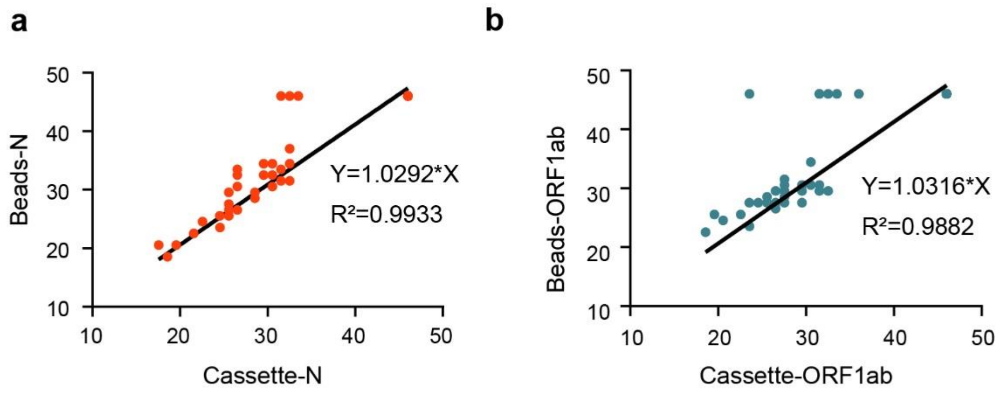


**Figure. S12 Comparison of CT values obtained through iDEP and conventional qPCR method. a,** Comparative analysis of CT values for the N gene detected using two distinct techniques. **b,** Comparative analysis of CT values for the ORF1ab detected using two distinct techniques.

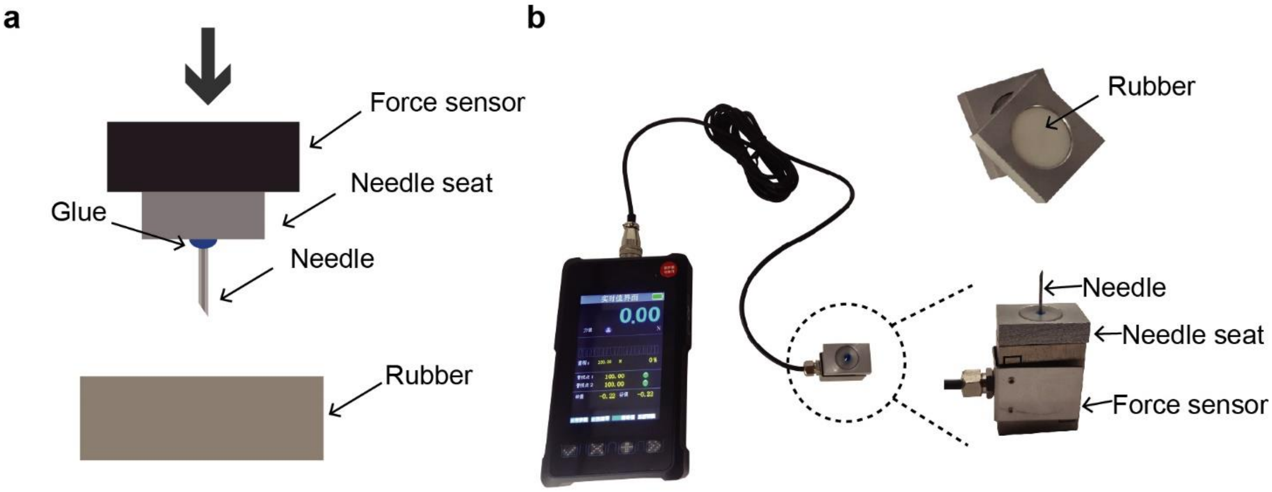


**Figure. S13 Force testing apparatus for puncturing rubber with a needle. a,** Schematic diagram of force testing for puncturing rubber. **b,** The photograph of the force sensor and a schematic diagram of the fixation of the hollow needle and rubber.

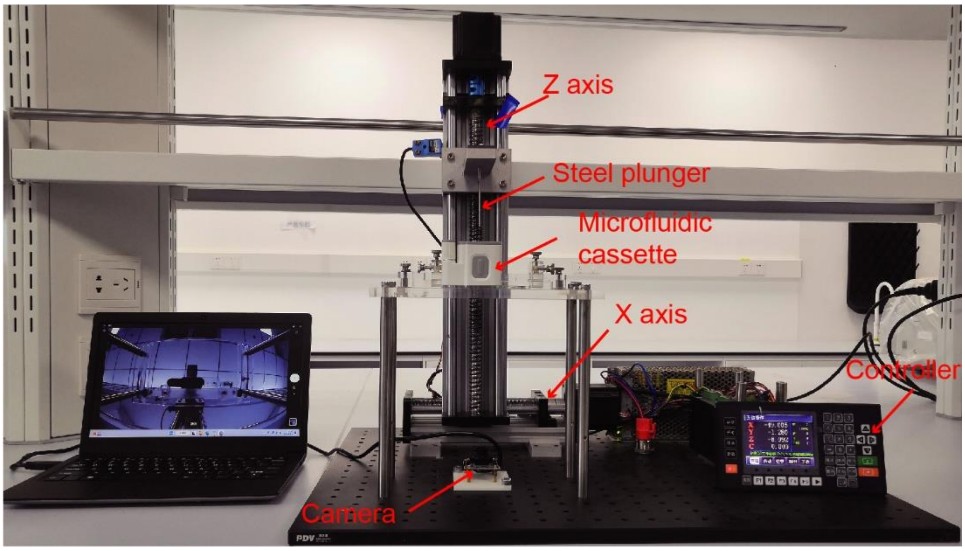


**Figure. S14 Photograph of the fluid testing platform.** The platform includes motion components in two directions, a motor controller, an image acquisition camera, and a laptop computer for display.
